# Mortality Trends for Cardiac Arrest with Acute Respiratory Failure Among U.S. Adults: A CDC WONDER Analysis From 1999–2023

**DOI:** 10.64898/2026.06.16.26355839

**Authors:** Dania Hussain, Habiba Nadeem Khan, Saif ur Rahman, Bilal Aslam, Anas Nasir, Muhammad Saad Bin Arshad Arain, Amna Zaman Khan, Maria Usman, Muhammad Saad Zahid, Haider Imran, Muhtasham Tahir, Rayyan Mohammad Makki Bakhsh, Areej Dar, Laiba Sultan, Sadia Ghafur, Muhammad Ali, Kamil Ahmad Kamil

**Affiliations:** United Medical and Dental College, Karachi, Pakistan; Central Park Medical College, Lahore, Pakistan; Bacha Khan Medical College, Mardan, Pakistan; University of Lahore, Pakistan; Sheikh Zayed Medical College, Rahim Yar Khan, Pakistan; Al-Aleem Medical College, University of Health Sciences, Lahore, Pakistan; Army Medical College, Rawalpindi, Pakistan; Foundation University Medical College, Islamabad, Pakistan; Sialkot Medical College, Sialkot, Pakistan; Shaikh Khalifa Bin Zayed Al-Nahyan Medical and Dental College, Lahore, Pakistan; Dow University of Health Sciences, Karachi, Pakistan; Dow International Medical College, Karachi, Sindh, Pakistan; Internal Medicine Department, Mirwais Regional Hospital, Kandahar, Afghanistan

## Abstract

**BACKGROUND:** Cardiac arrest(CA) and acute respiratory failure(ARF) are collectively at high risk of causing deaths among adults aged 25 and older in the United States. However, long-term trends to prevent these two coexisting conditions among adults are not well defined.

**OBJECTIVES:** The objective of this study was to analyse mortality trends for CA with ARF among U.S. adults aged 25 years and older from 1999 to 2023.

**METHODS:** Using the CDC WONDER Multiple Cause of Death database, we conducted a retrospective analysis of death certificates listing relevant ICD-10 codes for CA (I46) and ARF (J80, J96) among adults aged 25 years and older. Age-adjusted mortality rates (AAMRs) per 100,000 persons and the annual percentage change (APC) were calculated and stratified by demographics and geography. Trends were assessed using Joinpoint regression to estimate annual percentage change with 95% confidence intervals.

**RESULTS:** From 1999 to 2023, 807,236 deaths were recorded. The overall AAMR showed a significant upward trend (AAPC: 4.06%), rising sharply to a peak in 2021 (28.56) before declining. Males consistently had higher AAMRs than females. Both of them increased till 2021 and later decreased. Racial differences were observed in that Non-Hispanic (NH) Black individuals had the highest average AAMR, while NH Whites had the lowest. Geographically, the Western census region had the highest AAMR, increasing to 37.5 in 2021 (APC: 23.92; 95% CI: 16.21 to 28.35; p=0.0004), and rural areas demonstrated higher mortality than urban areas(13.45 vs 10.53). Adults aged 65 and older showed the highest AAMR, with a sudden rise to 96.7 in 2021 (APC: 17.4; 95% CI: 11.7 to 20.7, p<0.000001), followed by a subsequent decline, compared with the other age groups.

**CONCLUSIONS:** There was a marked AAMR due to CA and ARF over the past 24-year period, with a surge around the COVID-19 pandemic. Significant differences were observed by sex, race, and geography. These findings highlight that efforts are needed to prevent and manage mortalities by interventions among high-risk populations who have both HF and ARF.

## Introduction

Cardiac arrest (CA) refers to the sudden, unexpected stopping of the heart beating, often resulting in ischemia of vital organs. A complication requiring immediate intervention. On another note, acute respiratory failure results in decreased oxygen delivery to the body or the body’s inability to remove carbon dioxide from the lungs. This might result from a variety of causes, such as pneumonia, a severe asthma attack, or worsening chronic lung disease. When oxygen levels fall too low, or carbon dioxide builds up, it can place immense stress on the heart, sometimes causing it to arrest (1,2).

When a person experiences cardiac arrest or acute respiratory failure, time becomes the most precious and unforgiving factor. Every second of delay can worsen outcomes, and the window for meaningful intervention narrows rapidly. Not to forget, Cardiac arrest, in turn, cuts off blood flow to the lungs and every other organ, with lungs that may not recover immediately, leaving long-term complications (3).

These are both life-threatening emergencies in their own right, but when they occur together, which is being seen with increasing frequency in critically ill populations, the complexity of care and urgency of intervention rise dramatically (4,5). The United States experienced major trends in mortality due to CA, and research revealed substantial evidence associating CA with previous respiratory diseases and their interlinked relationship.

The interplay between the two conditions is particularly dangerous because one can so easily trigger or worsen the other. For instance, cardiac arrest leads to systemic hypoperfusion, exacerbating lung injury and increasing the risk of acute respiratory distress syndrome (ARDS) upon resuscitation (6). Conversely, respiratory failure causes severe hypoxia and acidosis, which can precipitate arrhythmias and, in some cases, full cardiac arrest by overwhelming the cardiovascular system (7). This bidirectional dynamic creates a vicious cycle, a downward spiral where each condition feeds into the other, making stabilisation and recovery significantly more difficult.

To address this gap, we thoroughly analysed national mortality data from the Centres for Disease Control and Prevention (CDC) between 1999 and 2023. This study aims to characterise mortality trends involving cardiac arrest and acute respiratory failure as co-occurring causes of death, using death certificate data from the CDC’s platform. By examining changes over time, we hope to contribute valuable insights into the evolving burden of this dual condition and inform future research, clinical decision-making, and health policy.

## METHODS

### Study design

The data for this descriptive analysis were retrieved using the Centres for Disease Control and Prevention Wide-Ranging Online Data for Epidemiologic Research (CDC WONDER) Database [6]. The International Classification of Diseases (ICD-10) codes I46 for Cardiac Arrest, J96 for Acute Respiratory Failure, J96.9 for Respiratory Failure, and J80 for Adult respiratory distress syndrome were used to extract data from death certificates [7]. This analysis included all individuals aged 25 and older who had Cardiac Arrest and Acute Respiratory Failure or Respiratory Failure or Adult Respiratory Syndrome listed anywhere on their death certificate in the US between 1999 and 2023. Data were retrieved from the multiple cause of death dataset. As CDC WONDER provides de-identified, publicly available data, Institutional Review Board (IRB) approval was not required. The Strengthening the Reporting of Observational Studies in Epidemiology (STROBE) guidelines were followed in this study [8].

### Data abstraction

Mortality data related to Cardiac Arrest and Acute Respiratory Failure for individuals aged 25 and older in the U.S. were collected from 1999 to 2023, except for urbanisation, for which data were extracted from 1999-2000. Data were stratified by year, gender, census region, race, urban-rural status, state, and place of death classification. Census regions—Northeast, Midwest, South, and West—were defined per the U.S. Census Bureau [6]. Urban and rural areas were classified using the 2013 National Centre for Health Statistics Urban-Rural Classification Scheme for Counties [9]. Areas with populations ≥50,000 were considered urban, while those <50,000 were categorised as rural. Counties were further divided into six levels of urbanisation: four metropolitan categories (large central metro, large fringe metro, medium metro, and small metro) were classified as urban, while two nonmetropolitan categories (micropolitan and noncore) were designated as rural. Racial and ethnic categories included non-Hispanic (NH) White, NH Black or African American, Hispanic or Latino, NH American Indian or Alaska Native, and NH Asian or Pacific Islander. Age-Adjusted Mortality rates (AAMR) per 100,000 persons were extracted with corresponding 95% confidence intervals (CI), standardised to the 2000 U.S. population.

### Statistical analysis

Temporal trends in age-adjusted mortality rates (AAMR) were evaluated using joinpoint regression to identify significant changes over time. To examine trends in Acute Respiratory Failure- and Cardiac Arrest-related mortality, the Joinpoint Regression Program (Version 5.3.0.0) was used [10]. This method fits linear segments on a logarithmic scale and detects “joinpoints” where trends shift [11]. As our research spans over 24 years, the software was configured to identify up to four joinpoints indicating significant trend changes. When trend variations were more distinct, fewer joinpoints were identified. Therefore, the analysis enabled detection of between 0 and 4 joinpoints. The Grid Search method (2, 2, 0), combined with empirical quantile and weighted Bayesian Information Criterion (BIC) methods, was applied to determine the optimal number of joinpoints. Annual percent changes (APCs) and their corresponding 95% confidence intervals (CIs) were calculated for each segment, while the average annual per cent change (AAPC) summarised the overall trend during the study period. APC and AAPC were deemed statistically significant if their 95% CIs excluded zero, with significance determined by two-tailed hypothesis testing at a threshold of p < 0.05.

## RESULTS

Between 2018 and 2025, there were 807,236 cardiac arrest and acute respiratory failure (ARF)-related deaths among U.S. adults, resulting in an overall age-adjusted mortality rate of 13.88 per 100,000. Men experienced a higher mortality burden compared to women, accounting for 51.53% of total deaths with a rate of 16.58 per 100,000. While Non-Hispanic (NH) White individuals comprised the greatest absolute number and proportion of deaths (549,910; 68.12%), the highest age-adjusted mortality rates were observed among NH Black or African American (21.81 per 100,000) and Hispanic/Latino (19.61 per 100,000) populations. Mortality increased substantially with age; adults aged 65 years and older represented 74.15% of all deaths and demonstrated a significantly elevated mortality rate of 52.98 per 100,000 (Table 1).

**Table 1:** Cardiac Arrest and ARF-Related Deaths, and Total Age-Adjusted Mortality Rates per 100,000 Among U.S. Older Adults, Stratified by Demographic Variables, 2018–2025.

| **Demographic Variable** | **Deaths** | **% of Total Deaths** | **Total Age-Adjusted**<br>**Mortality Rate (95% CI)** | **Population** |
| --- | --- | --- | --- | --- |
| **Overall** | 807,236 | 100% | 13.877 | 5,163,131,262 |
| **Men** | 415,948 | 51.53% | 16.576 | 2,491,041,251 |
| **Women** | 391,288 | 48.47% | 11.854 | 2,672,090,011 |
| **NH American Indian or Alaska Native** | 5,262 | 0.651% | 15.034 | 37,910,158 |
| **NH Asian** | 32,902 | 4.07% | 14.329 | 281,715,534 |
| **NH Black or African American** | 12,319 | 1.52% | 21.812 | 602,204,655 |
| **NH Native Hawaiian or Other Pacific Islander** |  |  |  |  |
| **NH White** | 549,910 | 68.12% | 12.28 | 3,528,912,956 |
| **NH More than one race** |  |  |  |  |
| **Hispanic/Latino** | 94,382 | 11.69% | 19.61 | 702,028,728 |
| **Adults Aged 25-44 Years** | 29,736 | 3.68% | 1.44 | 2,119,405,590 |
| **Adults Aged 45-64 Years** | 178,906 | 22.16% | 8.31 | 1,942,357,841 |
| **Adults Aged ≥ 65 Years** | 598,594 | 74.15% | 52.98 | 1,101,367,831 |

NH: non-Hispanic
\*Indicates total crude mortality rate rather than total age-adjusted mortality rate

### Overall

A total of 807,236 deaths were reported related to Cardiac Arrest with Acute Respiratory Failure. The AAMR increased non-significantly from 9.62 in 1999 to 9.80 in 2005 (APC: 0.27; 95% CI: -8.76 to 7.97; p=0.88), followed by a further increase to 16.73 in 2018 (APC: 4.06; 95% CI: -1.65 to 7.45). There was a significant increase to 28.56 till 2021 (APC: 19.62; 95% CI: 13.65 to 23.74; p<0.000001), and finally a significant decrease to 19.54 till 2023 (APC: -16.5; 95% CI: -23.7 to -8.96; p<0.000001). (Reference Supplementary Table 3)

### Gender

Males had a higher overall AAMR than females. The AAMR for males increased from 11.76 in 1999 to 11.59 in 2006 (APC: 0.32; 95% CI: -9.75 to 10.13; p= 0.909), followed by a further increase to 19.85 till 2018 (APC: 4.23; 95% CI: -2.07 to 8.22; p=0.112), followed by a significant increase to 35.05 till 2021 (APC: 21.25; 95% CI: 14.65 to 25.90; p<0.000001) and finally there was a significant decrease to 23.35 till 2023 (APC: -17.91; 95% CI: -25.56 to -9.82; p<0.000001). For females, the AAMR increased from 8.25 in 1999 to 23.19 in 2021, followed by a significant decrease to 16.54 till 2023 (APC: -14.64; 95% CI: -21.54 to -7.23; p<0.000001). (Reference Supplementary Table 1)

### Urbanization

When stratified by urbanisation, urban areas had a higher AAMR than rural areas (13.45 vs 10.53). The AAMR for urban areas increased from 10.27 in 1999 to 17.04 in 2018 (APC: 3.00; 95% CI: 1.94 to 3.73; p=0.008), followed by a significant increase to 24.05 in 2020 (APC: 19.26; 95% CI: 7.03 to 25.58; p<0.000001). The AAMR for rural areas increased from 6.84 in 1999 to 7.17 in 2005 (APC: 0.03; 95% CI: 10.23 to 4.54; p=0.909), followed by a further increase to 15.41in 2018 (APC: 6.42; 95% CI: 3.43 to 8.94; p=0.031), and finally a significant increase to 21.82 till 2020 (APC: 16.60; 95% CI: 7.06 to 21.58; p<0.000001). (Reference Supplementary Table 9)

### Race

From 1999 to 2018, all the different racial and ethnic groups in the United States diagnosed with cardiac arrest and acute respiratory failure showed a non-significant slow rise in their age-adjusted mortality rates (AAMR). However, a sudden spike was observed during the COVID-19 pandemic (2019-2021), followed by a decline from 2022 onwards. The most significant increase was seen in Non-Hispanic American Indian/Alaska Native individuals with their AAMR being the lowest among all in 1999 to a very steep rise till 19.21 in 2018 (p=0.002) and then a significant and sharp surge to 42.19 in 2021 (APC: 27.7; 95% CI: 17.01 to 34.02; p<0.000001) before decreasing to 21.5 in 2023 (APC: -28.39; 95% CI: -36.77 to -20.29; p<0.000001).

Other racial and ethnic groups also had a fair rise in mortality till 2018, followed by a significant increase during the COVID-19 period and then a decline afterwards.

Non-Hispanic Black/African American populations had their AAMR rise from 25.66 in 2018 (APC: 3.04; 95% CI: 2.3 to 3.6; p=0.0004) to 44.26 in 2021 (APC: 22.74; 95% CI: 16.28 to 26.5; p<0.000001).
Non-Hispanic White individuals saw AAMR increase from 15.2 (APC: 4.409; 95% CI: to 6.54; p=0.016) to 24.28 (APC: 16.11; 95% CI: 11.11 to 19.17; p<0.000001).
Non-Hispanic Asian/Pacific Islanders increased from 14.7 (APC: 0.23; 95% CI: -0.69 to 1.07; p=0.53) to 25.3 (APC: 24.08; 95% CI: 16.51 to 28.7; p<0.000001).
Hispanic/Latino individuals rose from 19.84 (APC: 1.23; 95% CI: -0.41 to 2.59; p=0.115) to 42.7 (APC: 31.98; 95% CI: 19.3 to 39.5; p<0.000001).

(Reference Supplementary Table 1 and 6)

## CENSUS DIVISIONS

AAMR for patients with cardiac arrest and acute respiratory failure increased very slowly across all the U.S regions (Northwest, Midwest, South, and West) from 1999 to 2018, followed by a notable peak in 2021 and a subsequent decline. The West had the highest mortality rate among all these geographical regions, with AAMR increasing to 37.5 in 2021 (APC: 23.92; 95% CI: 16.21 to 28.35; p=0.0004) from 13.4 in 1999 (APC: 2.14; 95% CI: 1.29 to 2.87; p=0.0004). In contrast, the Midwest showed the lowest AAMR, increasing from 4.6 in 1999 (p=0.657) to 18.4 in 2021 (APC:19.14; 95% CI: 14.7 to 21.7; p<0.000001). The Northeast experienced a steady increase from 12.0 in 1999 (APC: 2.13; 95% CI: 1.38 to 2.73; p=0.0004), peaking at 30.18 in 2020 (APC: 22.7; 95% CI: 15.06 to 27.9; p<0.000001), followed by a gradual decline through 2023. (Reference Supplementary Table 7)

## AGE GROUPS

Among all age groups, the highest mortality was seen among individuals aged 65 years and older. Their AAMR increased steadily from 39.2 in 1999 to 62.2 in 2018 (APC: 2.74; 95% CI: 2.05 to 3.33; p<0.000001) with a sudden rise to 96.7 in 2021 (APC: 17.4; 95% CI: 11.7 to 20.7, p<0.000001) followed by a subsequent decline. The lowest mortality trends were observed among younger adults (25-44 years), with AAMRs remaining around 1.0 through most of the study period and rising slightly to 4.5 in 2021 (APC: 37.15; 95% CI: 27.14 to 43.3; p<0.000001) during the COVID-19 pandemic. (Reference Supplementary Tables 3, 4 and 5)

## DISCUSSION

In this study, we evaluated the 24-year mortality data from the Centres for Disease Control and Prevention. We observed a number of substantial findings on cardiac arrest and acute respiratory failure-related mortality trends. The primary findings include: first, the overall age-adjusted mortality rate (AAMR) increased till 2021, which was followed by a decline afterwards till 2023; a sudden surge in mortality trends was observed from 2019 till 2021, primarily due to the COVID-19 pandemic; males predominantly had a higher AAMR in comparison to females. Second, among different racial backgrounds of the United States, NH Blacks had the highest mortality rates, followed by Hispanic or Latino, NH American Indian or Alaska Natives, NH Asian or Pacific Islander, and lastly, NH Whites had the lowest AAMR. Third, significant geographical and urban-rural discrepancies were observed with the Western region having the highest AAMR, followed by the Northeastern, Southern and Midwestern regions. Concurrently, rural areas exhibited a higher AAMR than urban areas.

Out-of-hospital cardiac arrest is associated with a 90% mortality rate, accompanied by a survival probability of less than 1 in 5 patients (12). Patients who experience cardiac arrest are at a high risk of acute respiratory failure (ARF). Respiratory failure is, in fact, the second most common organ failure associated with OHCA, mainly due to the increased susceptibility of the lungs to reperfusion injury and sepsis-like systemic inflammatory reaction. (13) Approximately 20% of ARDS cases are neglected, leading to suboptimal care and an increase in complications; furthermore, ARDS is associated with 40% in-hospital fatality rates due to subclinical diagnosis (14). Incidences and resultant mortality of CA and ARF increased during the COVID-19 pandemic, with a study reporting a twofold increased risk of CAD with COVID-19 (15). The recent decline in mortality trends can be attributed to several factors, including the initial decline in COVID-19 cases, improved healthcare management, increased use of non-invasive ventilation and LTVV, increased bystander CPR, therapeutic hypothermia, and use of 100% inspired oxygen therapy, followed by ROSC (16). The overall mortality rate is still alarmingly high; consequently, a need for further improvement in patient care to effectively manage the mortality rates and comorbidities persists.

Disparities among different racial groups across the United States were significant; the highest AAMR is exhibited by the Black population, followed by the Hispanic population, and the lowest is observed by the White population. The reason for this difference is multifactorial. According to a study, the rate of bystander CPR given to the Black and Hispanic populations, both at home and in public spaces, is considerably lower in comparison to their white counterparts (17). The use of automated external defibrillators is also low for the black population; a study has shown that the use of AED along with CPR considerably improved the survival rates in comparison to when the CPR was done alone (18). The overall usage of AEDs in the United States is inadequate. In 2019, the use of AED was limited to only 7%, even though the incidence of cardiac arrest with a shockable rhythm was 20%. (18) Moreover, despite residing close to high-end treatment centres, the Black and Hispanic populations are more likely to receive treatment in low-quality hospitals. In addition, the probable use of therapeutic hypothermia given after cardiac arrest to provide neuroprotection is low in the Black and Hispanic population (18). Some studies also highlighted the improbable use of carotid arteriogram among minority races. (18) Several studies previously reported lower rates of CVD among Hispanics despite living in low socioeconomic conditions, a term referred to as the Hispanic paradox. Our study significantly shows this contradictory finding of the higher mortality trends among Hispanics. The higher prevalence of DM and obesity, the direct risk factors of SCA, among Hispanics and Blacks could also be contributing to this disturbing trend (19).

Previous studies also display a significantly higher risk of mortality from acute respiratory failure among minorities in comparison to the ethnic majority. (20) Black patients, when evaluated by pulse oximetry, have shown a three-fold rise in occult hypoxemia in contrast to NH White individuals, and this was linked to higher mortality rates. (21) In addition, another study highlighted that among the ethnic minorities, the increased risk of occult hypoxemia was due to inadequate oxygen delivery. (21). Hispanic and Black populations have lower rates of insurance, which ultimately decreases the likelihood of receiving standard treatment for chronic diseases. (22) A decreased use of invasive ventilation in Black, Hispanic and Asian populations has been observed, which, in the setting of severe hypoxemia, caused an increase in 28-day mortality. Linguistic barriers in minorities are found to be correlated with unfavourable outcomes for individuals with chronic diseases in the community health centres (20). A study that modified the socioeconomic factors found an appreciable reduction in health disparities among the minority group, accentuating the significance of targeted interventions to promote health equity. (21)

Males exhibited higher SCA and ARF-related mortality throughout the study period in comparison to women. Although the exact cause for this difference is not fully known, females have a low incidence of cardiac arrest, likely influenced by the cardioprotective effects of estrogen and progesterone. In contrast, men experience higher incidences of both OHCA and IHCA (23, 24). However, it is notable that females have lower incidences of bystander CPR and initial shockable rhythms and are less likely to undergo evidence-informed intervention such as PCI and therapeutic hypothermia, leading to a low post-discharge survival (25). Males develop CAD at an earlier age and are at a high risk of experiencing hypertensive emergencies, which can potentially lead to comorbidities such as CKD (26). Hospital admission rates and post-discharge survival rates are higher for men. Another notable difference is the higher incidence of the favourable tachycardias VF/VT among men in comparison to women, causing lower survival rates in women (27)

On the other hand, women are at high risk of developing ARF owing to the immunological effects of estrogen, which can lead to increased inflammatory response and severity of disease; despite this, women are less likely to receive invasive procedures for ARF in comparison to men (28). One study reported no significant differences in overall mortality or the need for invasive ventilation between genders (29). Additional studies are necessary to understand the exact causes of this disturbing trend, and implementing more targeted interventions is important to address the disparities more efficiently.

The mortality trend in individuals aged 65 and above is 7 times higher than in the 45-64 age group. IHCA and OHCA are both associated with adverse outcomes in this patient population, as they exhibit multiple comorbidities with IHD, functional impairment, and increased susceptibility to risk factors (30) (31). Moreover, this patient group is disparately treated with IMV, receives less aggressive care as the degree of intervention scales back with increasing age, and the frequency of discontinuing critical care or advanced life support is high in older patients. (32).

The mortality trend was highest in the Western region, which comprises a large Hispanic population, and lowest in the Midwestern region. Individuals residing in low-income areas are less likely to receive bystander CPR than those living in high-income areas (33). Low socioeconomic status is associated with limited access to specialised health care, targeted interventions and decreased rates of follow-up for chronic diseases. Moreover, states without expanded Medicaid coverage, consisting mostly of Hispanic and Black populations, have higher rates of cardiovascular mortality than the states with expanded Medicaid coverage (34). The urban areas exhibited a higher mortality trend than rural areas, likely due to increased population concentration and spatial delays in treatment, which are more pronounced in urban areas and higher in rural areas due to a lack of efficient transportation. The rate of bystander CPR is higher in rural areas, while the rate of defibrillation is lower (35). Economically disadvantaged populations are also at a higher risk of OHCA. Rural areas also have a higher incidence of ARF and are less likely to receive appropriate treatment, causing increased mortality rates in comparison to urban areas (36). Directing public health initiatives, including enhancing the specialised healthcare availability, advanced cardiopulmonary resuscitation education, prompt diagnosis of ARF, optimal post CA care, medical infrastructure optimization, promoting health education outreach efforts, and advocating lifestyle measures among the affected population, is an immediate necessity to mitigate the impact of this condition, to substantially lower the disparities and to improve the overall mortality (37).

## LIMITATIONS

We acknowledge that our study comprises several limitations. First, reliance on ICD-10 codes might lead to discrepancies or overlooking cardiac arrest and acute respiratory failure as a cause of death. Second, the omission of critical insights on the CDC database, such as lab data analysis, genetic analysis and the baseline characteristics involving unresponsiveness, absent pulse and respiration, hypoxemia, cyanosis, altered mental status, abnormal immune function and biomarkers, is not available, which limits the precision of our findings. Third, there is the absence of data about medical therapy and treatment of Cardiac arrest and acute respiratory failure. Lastly, socioeconomic factors affecting medical access that might reconfigure the outcomes are not available in the database.

## Conclusion

Between 1999 and 2023, age-adjusted mortality rates (AAMRs) due to cardiac arrest and acute respiratory failure showed a gradual rise across all racial, age, and regional groups, with a sharp increase during the COVID-19 pandemic (2019–2021) followed by a decline post-2022. The most dramatic increase was observed among Non-Hispanic American Indian/Alaska Native individuals and residents of the Western U.S. Older adults (≥65 years) consistently showed the highest mortality, while younger adults showed a relatively low but significant spike during the pandemic period. These findings show that the COVID-19 pandemic affected some communities more than others, especially certain racial and regional groups. This suggests a need for better access to healthcare, earlier detection, and improved critical care to prevent these serious outcomes.

## Data Availability

The data underlying this article are publicly available in the Centers for Disease Control and Prevention Wide-Ranging Online Data for Epidemiologic Research (CDC WONDER) Multiple Cause of Death database, accessible at https://wonder.cdc.gov/.

## Funding Declaration

The authors received no financial support for the research, authorship, and/or publication of this article.

## Acknowledgments/Disclosures

A portion of this study’s preliminary results was presented at the American Heart Association (AHA) Scientific Sessions 2025, November 8–10, 2025, and published as a meeting abstract in *Circulation* (Abstract Sat1402).

---

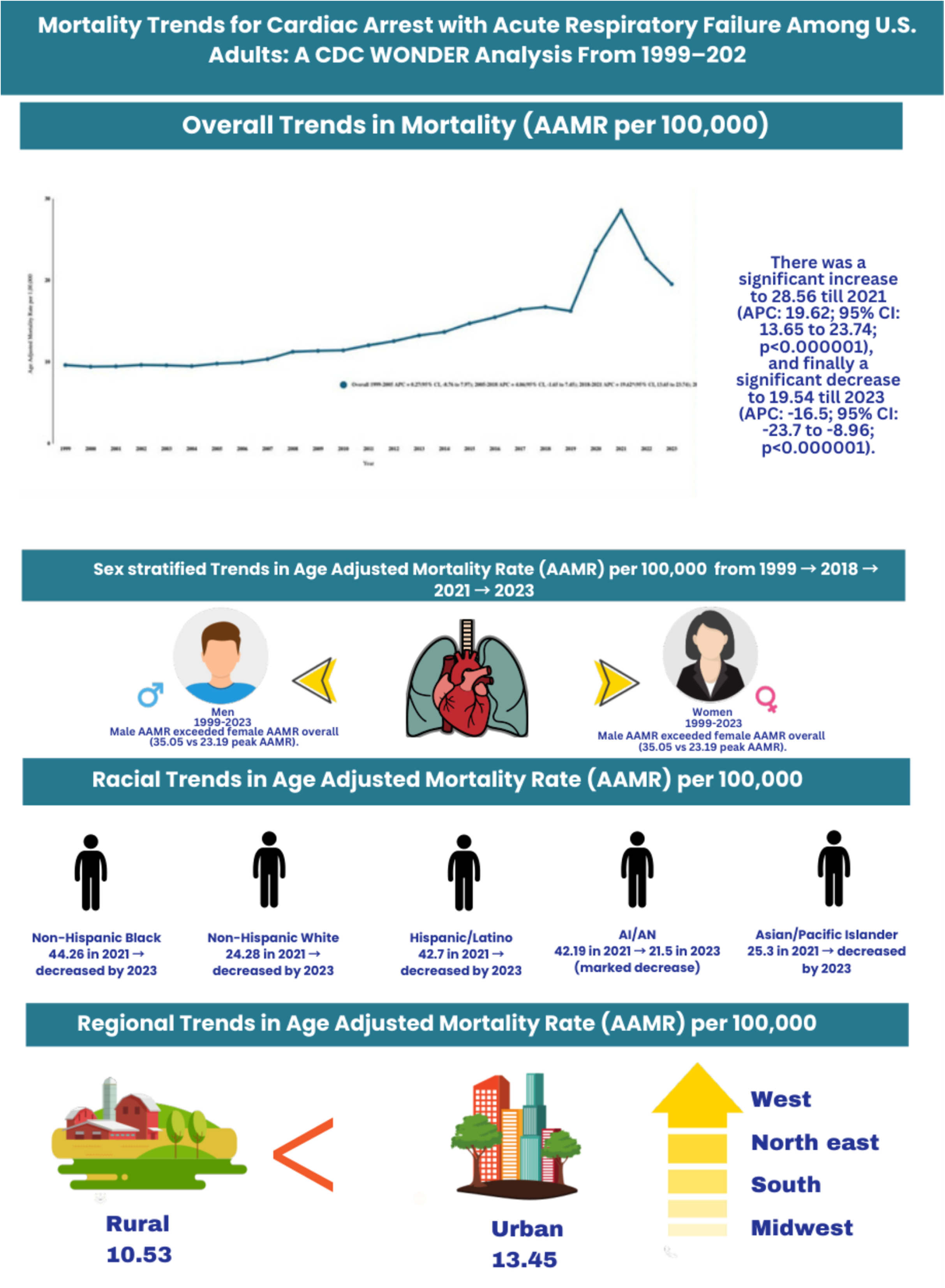

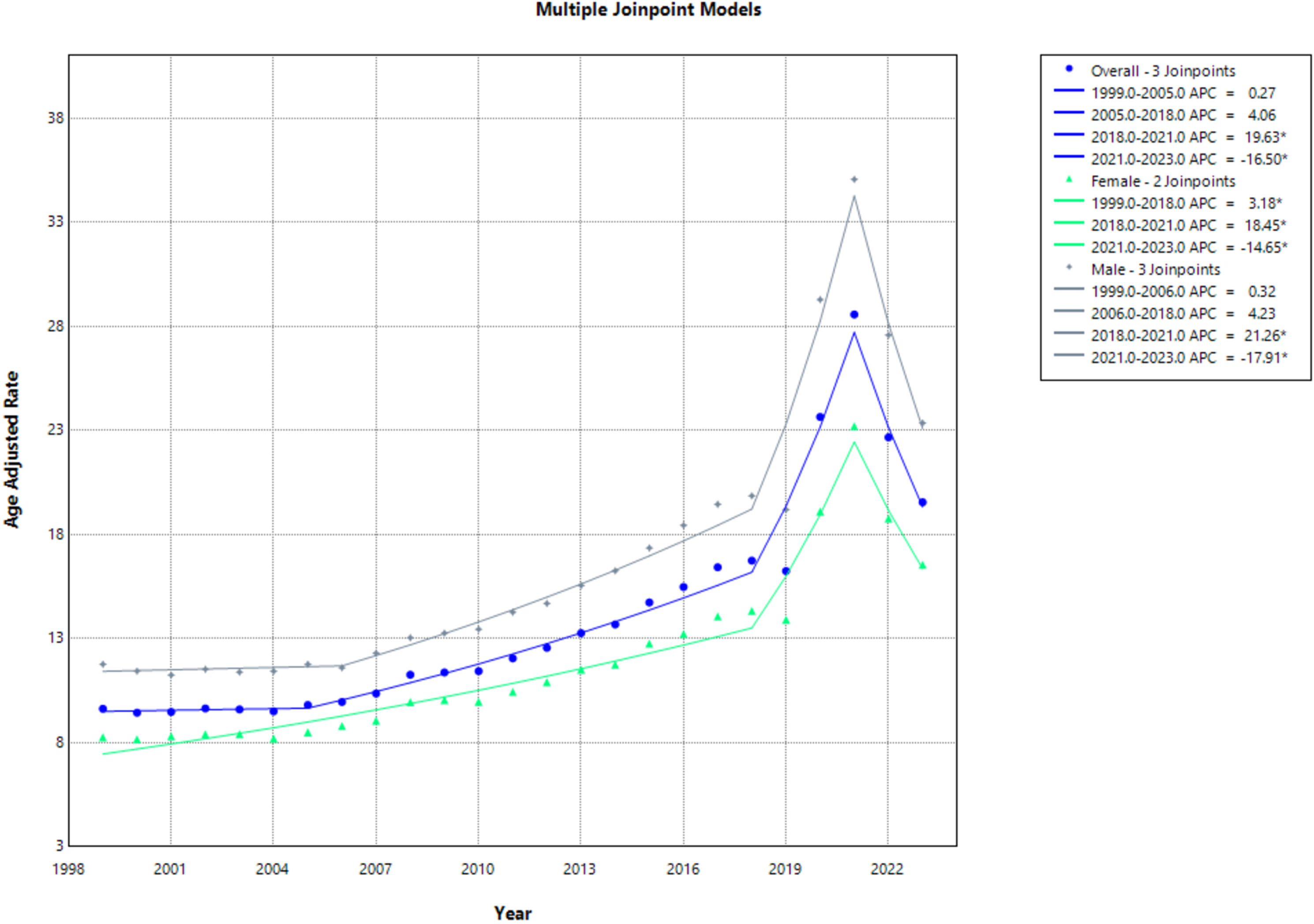

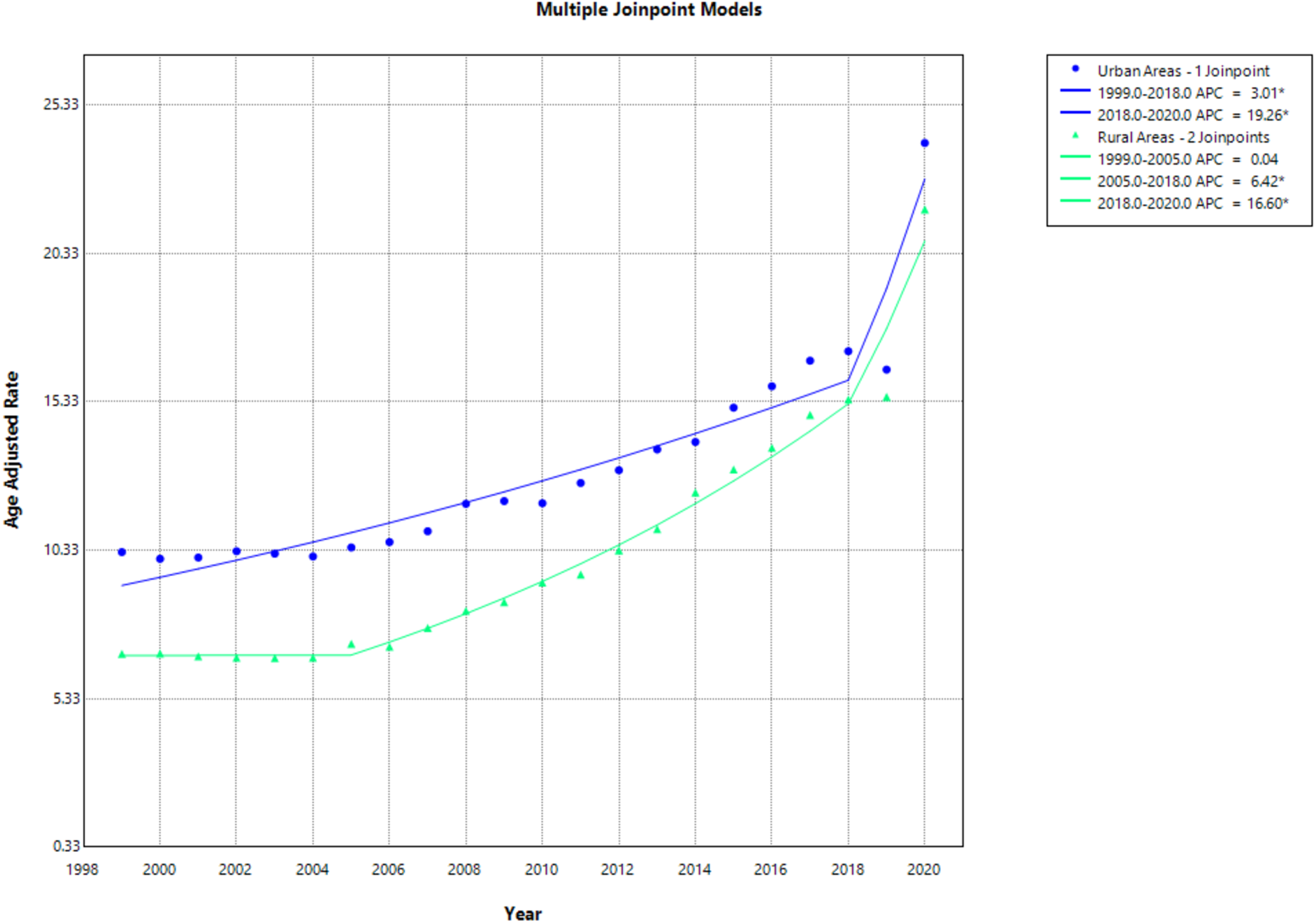

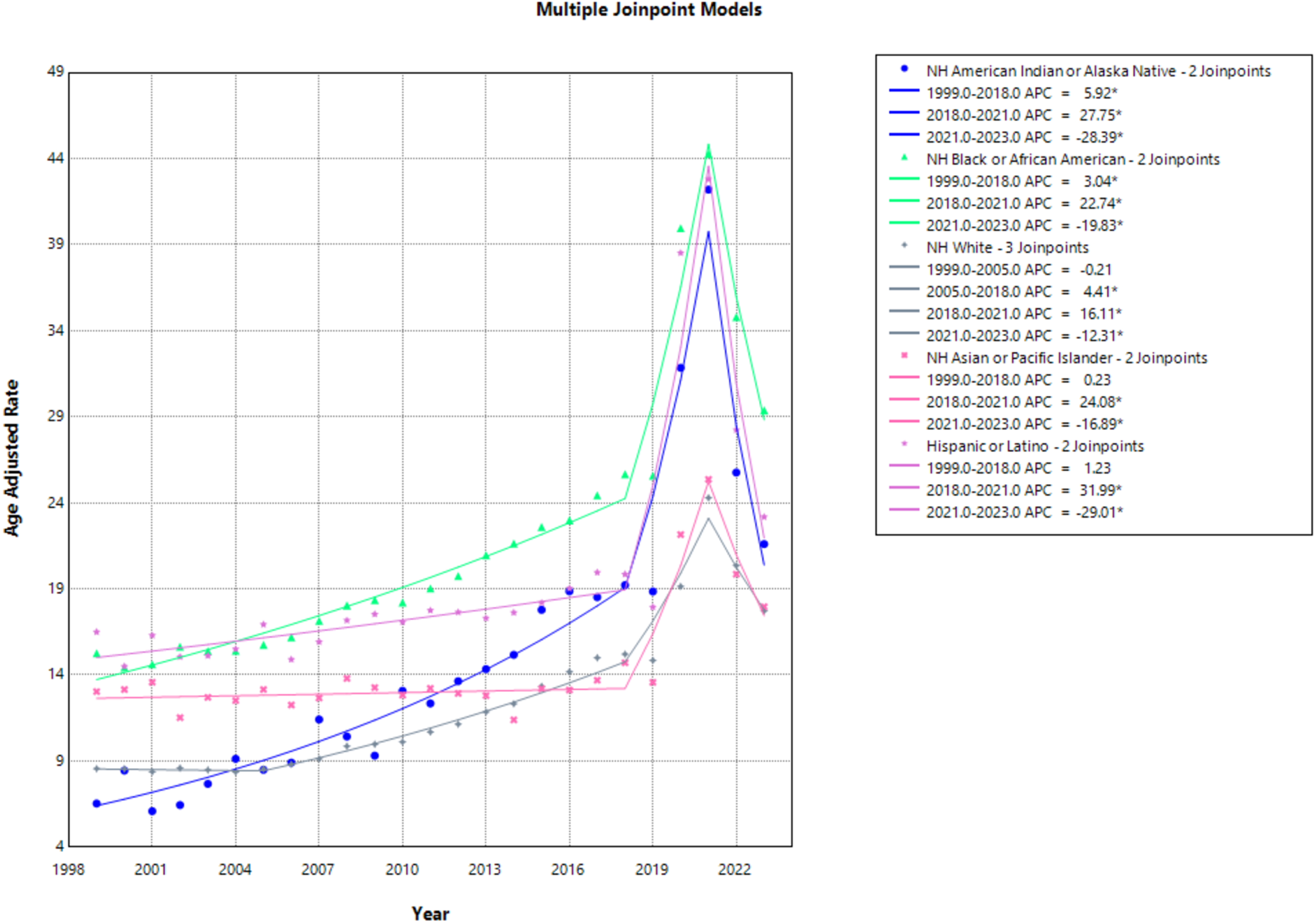

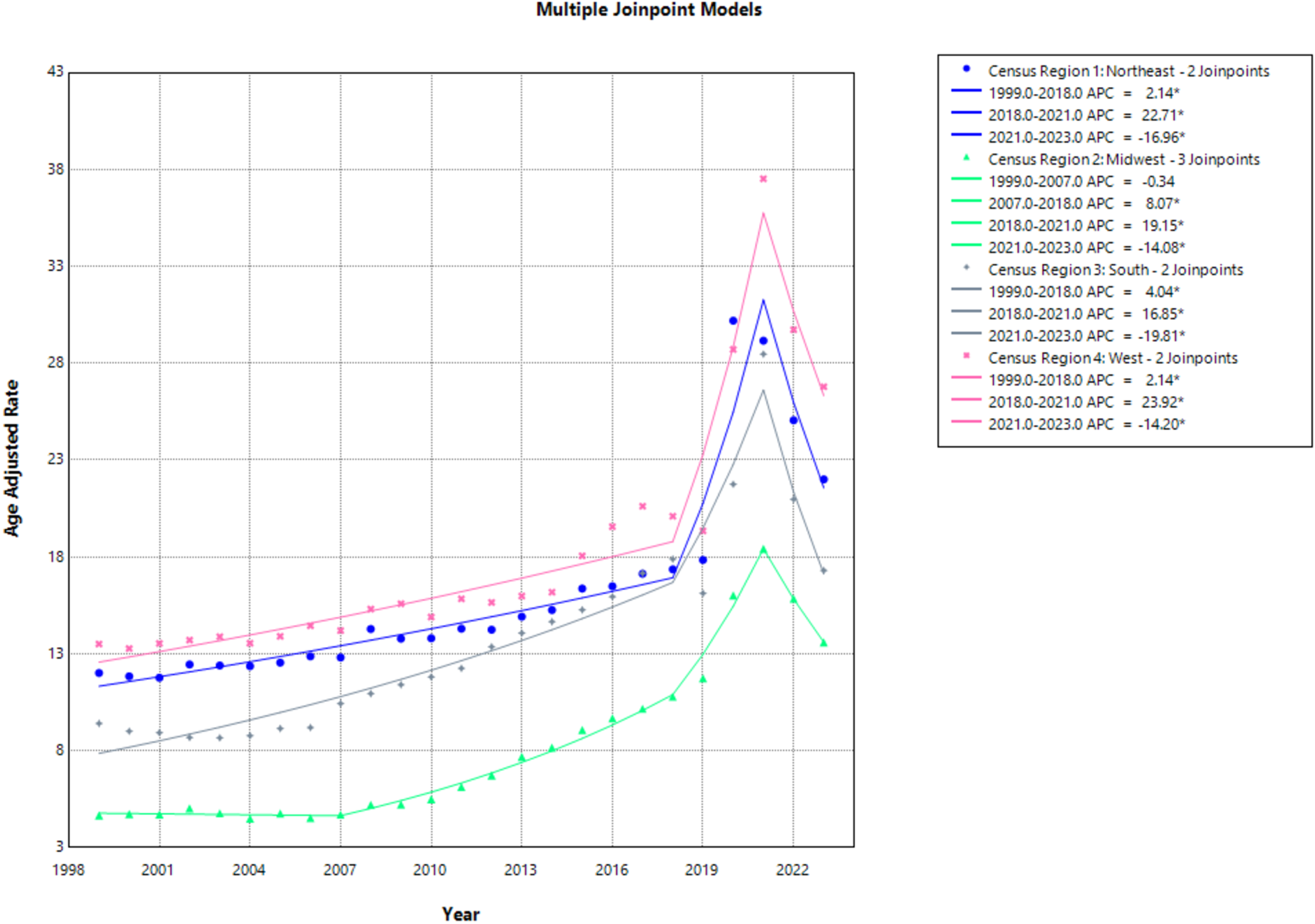

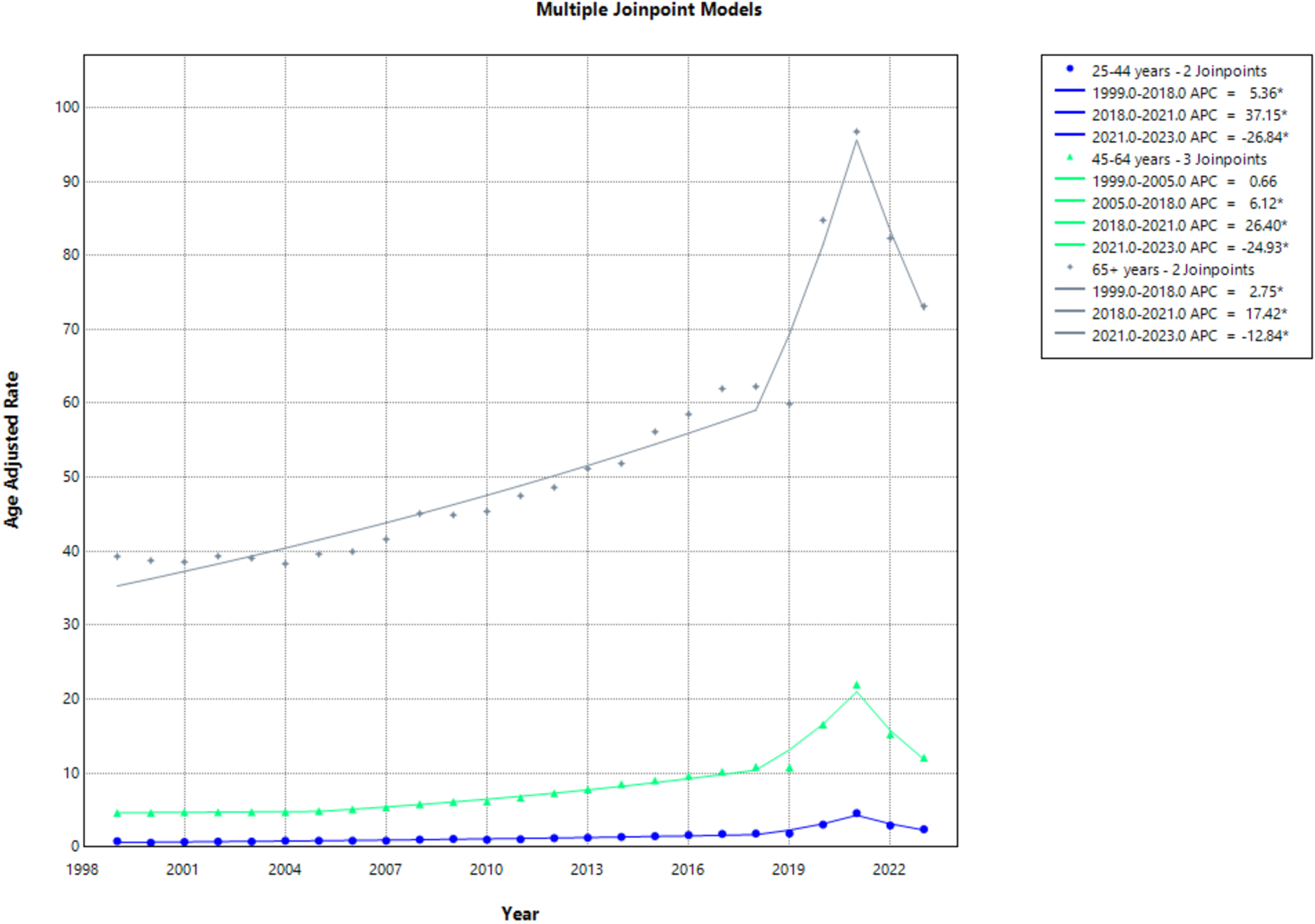

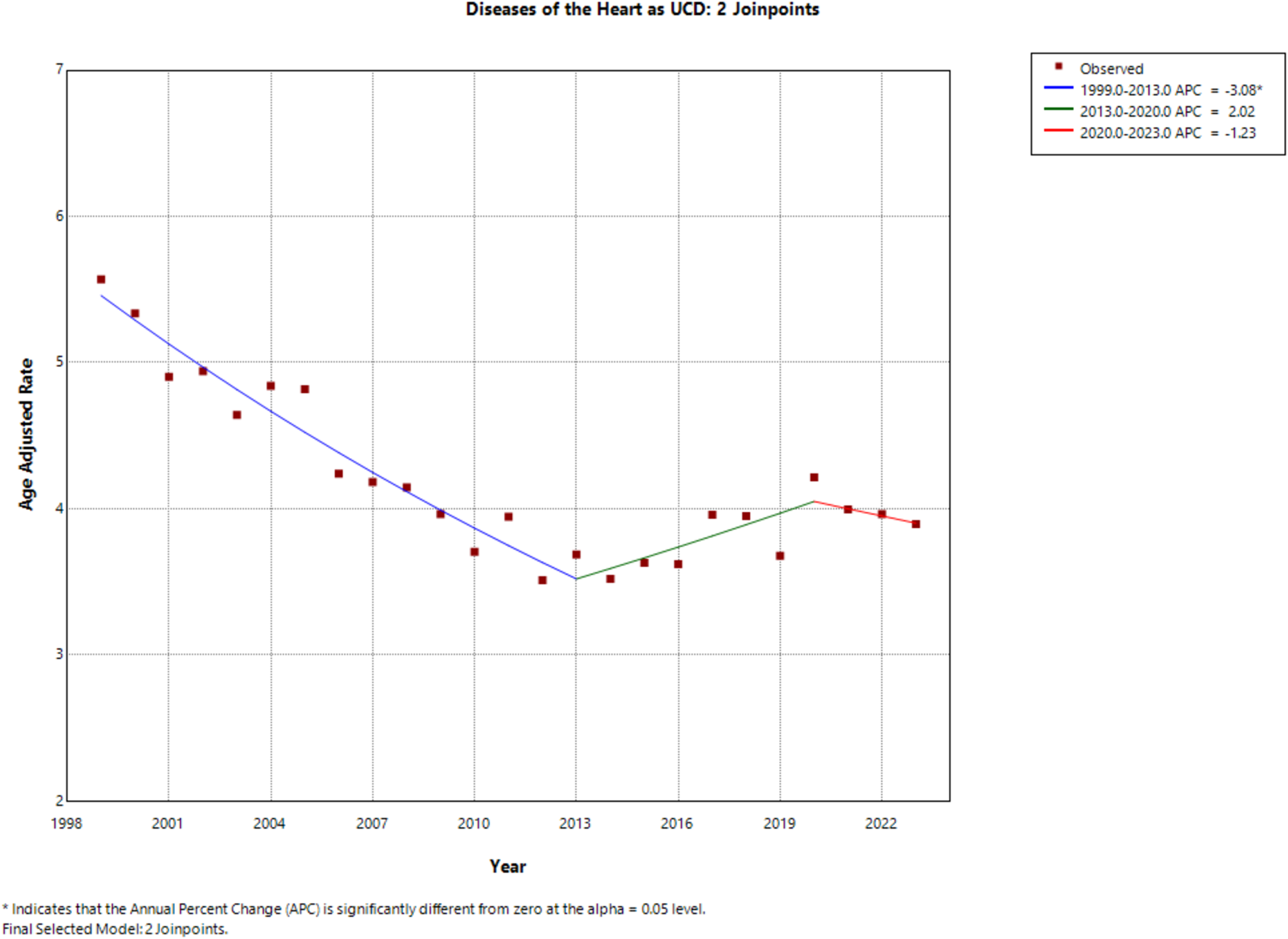

## Notes

### Competing Interest Statement

The authors have declared no competing interest.

### Clinical Trial

Not applicable. This study is a retrospective observational analysis of a publicly available, fully de-identified national database (CDC WONDER) and does not involve any prospective clinical trials or interventions.

### Funding Statement

The authors and their respective institutions received no external funding, financial support, or grants for any aspect of this submitted work. Furthermore, no payments or services were received from any third party for study design, data collection and analysis, manuscript preparation, or statistical analysis. The authors declare no external financial support was involved in the research, authorship, and/or publication of this article.

### Author Declarations

This study utilized publicly available, fully de-identified mortality data from the Centers for Disease Control and Prevention (CDC) WONDER database. In accordance with standard ethical guidelines for public database research, this study was deemed exempt from Institutional Review Board (IRB) review and patient informed consent requirements.

